# Do low TB prevalence and lack of BCG Vaccinations Contribute to Emergence of Multisystem Inflammatory Syndrome in Children?

**DOI:** 10.1101/2020.07.18.20156893

**Authors:** Tareef Fadhil Raham

## Abstract

**Background:** Emergence of new multisystem inflammatory syndrome in children (MIS-C) is thought to be associated with COVID-19 pandemic. Covid-19 morbidity and mortality variances among countries have been suggested by previous works to be influenced by BCG and previous latent TB infection (which is reflected by TB prevalence) possibly through inducing heterogeneous immunity against SARS-CoV-2.

**Aim:** To examine influence of BCG status and TB prevalence on variances among countries which report new multisystem inflammatory syndrome in children (MIS-C).

**Methods:** We choose all countries which report MIS-C till 23/6/2020, number of cases for each 10 million inhabitants was examined among 3 categories of countries classified according to BCG program status. TB prevalence, MIS-C no. / 10 million (M) population and Covid-19 deaths/M are taken as markers. Receiver operation characteristic - (ROC) curve, with some relative indicators such as (sensitivity and specificity rates), estimation area of trade - off between sensitivity and specificity, and cutoff points are used with different studied markers for discriminating different three pairs of countries (which have different BCG practices).

**Results:** BCG vaccinations and high TB prevalence are found to be associated with decrease MIS-C no. and COVID-19 deaths

**Conclusions:** Findings might explain variances in MIS-C incidence and in COVID-19 mortality among countries worldwide. Further studies to confirm this relation and to confirm possible similar relations in Kawasaki disease(KD) in previous epidemics is recommended.

**What is Known:**

- Although the etiology for KD remains unknown, available evidence supports the hypothesis that the pathogenesis is closely associated with dysregulation of immune responses to an infectious agent.
- BCG and / or Latent TB have heterogeneous beneficial effects.

**What is New:** Our study shows that TB prevalence and implementing BCG vaccination have negative statistical association with MIS-C cases and COVID-19 mortality.

## Introduction

In April 2020, reports emerged from the United Kingdom of a presentations in children similar to incomplete Kawasaki disease (KD) or toxic shock syndrome (TSS)^1,2,3^. In Italy, approximately 10 suspected Kawasaki-like disease cases have been recorded since 1 January 2020, eight of which were reclassified as new pediatric inflammatory multisystem syndrome temporally associated with SARS-CoV-2 infection in children (PIMS-TS)^4^ or multisystem inflammatory syndrome in children (MIS-C) or pediatric hyper inflammatory syndrome, or pediatric hyper inflammatory shock^1^which is hyper inflammatory syndrome with multiorgan involvement have some features similar to those of Kawasaki disease and TSS^5^. MIS-C cases also been reported since then in USA and countries in Europe. Classical KD with concurrent COVID-19 also have been reported^6^.

(KD) is an acute febrile systemic vasculitis that predominantly occurs in children younger than 5 years of age and the most common acquired heart disease during childhood in most industrialized countries^7^. The annual incidence of KD is highest in Japan, with more than 300 per 1000000 children aged 4 years or younger affected^8^, compared with 25 per 1000000 children aged 5 years or younger in North America^9^and is rarely reported in sub-Saharan Africa^10^. Although KD is less common in western countries KD shock syndrome (KDSS) a rare form of KD has a higher incidence in these countries (2.60 to 6.95%)^11,12, 13, 14^, compared with Asian countries like Taiwan which reported lower incidence rate (1.45%)^15^.

KDSS is often associated with myocarditis and requires critical care support during the acute phase of illness KDSS might mimic toxic shock syndrome^11^. The severity of inflammation in KD is reflected by inflammatory parameters; thus, laboratory findings are helpful for diagnosing incomplete KD just like MIS-C. Patients are considered to have incomplete KD when they lack a sufficient number of criteria to fulfill the epidemiologic case definition^1^. “Atypical KD” should be used in the presence of an unusual or odd manifestation of KD (e.g., nephritis or central nervous system complication)^16^.

A high level of circulating pro-inflammatory cytokines might contribute to the distributive component of shock. Indeed, KDD has been found associated with high levels of IL-6, and procalicitonin^12^. Patient with MIS-C also high levels of inflammatory markers, including procalcitonin, and serum interleukin 6^17^. Furthermore elevation of inflammatory markers (cytokines) has been previously demonstrated in inflammatory state for multiple conditions^18^including cytokine storms. Cytokine storm denotes a hyperactive immune response characterized by the release of interferons, interleukins, tumor-necrosis factors, chemokines, and several other mediators. Cytokine storms can be caused by a number of infectious, especially viral respiratory infections such as H5N1,H1N1 influenze and SARS-CoV2^19,20^ Other causative agents include the Epstein-Barr virus, cytomegalovirus^21^. Likewise in KD circumstantial evidence points to an infectious cause^22^. In the past 20 years, viruses of the coronavirus family have been proposed as possibly implicated in the pathogenesis of Kawasaki disease in small percentage^23,24^.

The case definition of MIS-C was regarded by some experts as quite broad and overlaps with Kawasaki disease,^25^. On the contrary other experts are concerned that current diagnostic criteria may not capture the true scope of the problem^26^.

Erythema or induration of the Bacillus Calmette–Guérin (BCG) injection site has been listed by the American Heart Association as a significant finding among the diagnostic guidelines for KD^27^. Such finding might account to 50%of patients^28,29^ BCG reactivation is thought to happen due to suggested cross reactivity of mycobacterial Heat Shock Protein 65 (HSP65) and the human homologue HSP63. Furthermore, cross immune heterogenecity was suggested between BCG and certain viral diseases since BCG induces trained immunity through expressing heterologous antigens of different pathogens which have been developed and tested since 1991^30^. Different studies have already show statistical associations of BCG vaccinations and TB prevalence status with reduced severity of COVID-19 disease^31,32,33,34, 35^. These studies suggest that BCG and natural latent TB infection induce heterogeneous immunity against SARS-CoV2. Studies are lacking regarding possible role of BCG or possible latent TB protective effects on incidence of MIS-C or KD including KDSS. This study is the first in aspect of looking for MIS-C incidence in relation to TB prevalence status and COVID-19 deaths in different countries classified according to BCG vaccination status and will address possible cause of disparities raised regarding variances in incidence of MIS-C and COVID-19 deaths rates among countries.

This study also discusses possible relation of KD with previous pandemics according to various TB prevalence and BCG status of some countries.

## Material and Methods

Data regarding to MIS-C cases collected till 23/6/2020. Cases outside this date are not included. The total number of countries is 15 countries. 13 country were listed by ECDC rapid risk assessment report on May, 15, 23020 as the only countries in the world which have been reported MIS-C cases, both Russia and Peru reported MIS-C cases on June,17 and June, 22 respectively are included in the study . These data are publically available through:

1. -Reference for data for all countries except (USA, Russia and Peru): European Centre for Disease Prevention and Control. Rapid Risk assessment. Paediatric inflammatory multisystem syndrome and SARS-CoV-2 infection in children – 15 May 2020. ECDC: Stockholm; 2020. https://www.ecdc.europa.eu/sites/default/files/documents/covid-19-risk-assessment-paediatric-inflammatory-multisystem-syndrome-15-May-2020.pdf
2. -Reference for data for Russia: Семенова, Мария (17 June 2020). “В Москве умер первый ребенок из-за новой болезни, вызванной COVID-19” (in Russian). RIA Novosti. Retrieved 23 June 2020.
3. -Reference data for Peru: Yáñez JA, Alvarez-Risco A, Delgado-Zegarra J (June 2020). “COVID-19 in Peru: from supervised walks for children to the first case of Kawasaki-like syndrome”. *BMJ*. 369: m2418. doi:10.1136/bmj.m2418. PMID 32571770. S2CID 219970740.
4. -Reference data for USA: We consider the following reference: Feldstein LR, Rose EB, Horwitz SM, et al. Multisystem Inflammatory Syndrome in U.S. Children and Adolescents. *N Engl J Med*. 2020;383(4):334-346. doi:10.1056/NEJMoa2021680 https://www.nejm.org/doi/full/10.1056/NEJMoa2021680

According to this, we consider 186 cases reported in *N Engl J Med* accounting cases for all states rather than 85 cases reported by ECDC rapid risk assessment report which account cases just for New York.

European Centre for Disease Prevention and Control (ECDC) reported 125 cases in France, while national surveillance reported 108 cases^36^.

We consider data of ECDC rapid risk assessment report so as to avoid bias within European countries.

Furtherly we check Google for any possible cases up to 23/6/2020. We look for any data whether population-based, cross-sectional studies, surveillances, news and others. The check includes all countries across the world in addition checking already included 15 countries for any extra reported or missed cases.

We check also: Wikipedia. Multisystem inflammatory syndrome in children https://en.wikipedia.org/wiki/Multisystem_inflammatory_syndrome_in_children#cite_note-

We consider COVID-19 deaths/Million as it is 2/7/2020 considering lag time of 10 days between day of case report and death^37,38^. (Appendix A)

Appendices B & C show countries’ TB prevalence and BCG practices data and references We put these countries in three different groups:

1. - No vaccination at all and no previous BCG: USA and Italy.
2. - No current vaccination but with previous vaccination program (countries which ceased BCG vaccinations previously): 9 countries: Austria, Canada, France, Germany, Luxembourg, Spain, Sweden, Switzerland, United Kingdom.
3. - Currently giving mass BCG vaccine through national program: Greece, Portugal, Peru, Russian Federation

Total countries were 15, they were distributed among that groups status as shown in table No. (1).

**Table No. (1):** *MIS*-C No./10M, COVID -19 deaths/Million versus BCG practice group of countries

| Countries | MIS cases /10M. pop. | Year Stopped mass BCG vaccination | COVID deaths /M. | TB Prevalence |
| --- | --- | --- | --- | --- |
| **1-No vaccination at all no previous BCG** |  |  |  |  |
| Canada | 3.179 | 0/0 | 228 | 6 |
| Italy | 1.654 | 0/0 | 575 | 7 |
| United State | 5.619 | 0/0 | 395 | 3 |
| Mean | 3.484 | - | 399.3 | 5.3 |
| **2-No current vaccination (previous vaccination)** |  |  |  |  |
| Austria | 1.110 | 1990 | 78 | 7 |
| France | 19.15 | 2007 | 457 | 9 |
| Germany | 0.596 | 1998 | 108 | 7 |
| Luxembourg | 79.875 | 2005 | 176 | 8 |
| Spain | 4.705 | 1981 | 607 | 9 |
| Sweden | 2.971 | 1975 | 532 | 6 |
| Switzerland | 3.466 | 1987 | 227 | 6 |
| United kingdom | 5.892 | 2005 | 647 | 8 |
| Mean | 14.29 | - | 354 | 7.5 |
| **3-Currently giving BCG vaccine** |  |  |  |  |
| Greece | 0.959 | Not stopped | 18 | 5 |
| Portugal | 0.981 | Not stopped | 155 | 24 |
| Peru | 0.303 | Not stopped | 299 | 123 |
| Russian Federation | 0.891 | Not stopped | 66 | 54 |
| Mean | 0.78 | - | 134.5 | 51.5 |

COVID -19 mortality, data TB prevalence data, and population data are obtained from publically published data. BCG data are obtained from publically available WHO data and various references.

Receiver operation characteristic - (ROC) curve, as well as some relative indicators such as (sensitivity and specificity rates), and cutoff points are used for discriminating different three categories of countries (which have different BCG practices). Discrimination of these categories done through studied markers. All statistical operations were performed through using the ready-made statistical package SPSS, ver. 22.

Table 1 shows collected data for the study sample in terms of country no. of MIS-C cases, BCG status, COVID -19 deaths and TB prevalence.

Patient and public involvement statement:

It was not appropriate or possible to involve patients or the public in this work given that we used general practice level summated data and related publically published morbidity and mortality statistics. Patients were not involved

### Results and Findings

Group 1 (countries with no vaccination at all and no previous BCG national programs) shows highest mean value for COVID-19 deaths/M inhabitant and lowest for TB prevalence.

Group 3 (countries currently giving BCG vaccine) shows lowest MIS-C no./10 M and lowest COVID-19 mortality and highest TB prevalence.

Group 2 No current vaccination (previous vaccination) group shows highest MIS-C and other parameters were in between.

Table No. (2) and fig (1) show estimated area of trade - off between sensitivity and specificity of markers through plotting sensitivity against a complement specificity outcome to examine the trade - off, which is called (ROC) curve, table 2 also show a significant level for testing area under the guideline of fifty percent, with 95% confidence interval of all probable combinations of pairs of countries and were as follows:

1- Pair of countries with no vaccination program at all (x axis) & no current vaccination program but have previous vaccination program (y axis): The common think in this pair is that both groups had no vaccination programs at this time whether had previous history of vaccinations or not.

2- Pair of countries with never gave BCG vaccination program (x axis) & those with currently giving BCG vaccination (y axis).

3-Pair of countries with currently no vaccination (ceased vaccination program) (x axis) & currently vaccinated (y axis). The common think in this pair is having vaccination whether in the past or at the current time.

**Table: (2):** Relationships among a diseased groups & controlled for all probable combinations of Disclusion time test in different parameters

| Group's Combinations | Parameters | Area | Std. Error | Asymp. Sig. | Asymp. 95% C.I. |  |
| --- | --- | --- | --- | --- | --- | --- |
|  |  |  |  |  | L.b. | U.b. |
| **I X II** | MIS cases /10M pop. | 0.583 | 0.175 | 0.683 | 0.241 | 0.926 |
|  | COVID-19 Deaths /M. | 0.417 | 0.173 | 0.683 | 0.078 | 0.755 |
|  | TB Prevalence | 0.833 | 0.133 | 0.102 | 0.572 | 1.094 |
| **I X III** | MIS cases /10M pop. | 0.000 | 0.000 | 0.034 | 0.000 | 0.000 |
|  | COVID-19 Deaths /M. | 0.083 | 0.115 | 0.077 | 0.000 | 0.309 |
|  | TB Prevalence | 0.833 | 0.170 | 0.157 | 0.500 | 1.167 |
| **II X III** | MIS cases /10M pop. | 0.094 | 0.095 | 0.027 | 0.000 | 0.280 |
|  | COVID -19 Deaths /M. | 0.188 | 0.135 | 0.089 | 0.000 | 0.452 |
|  | TB Prevalence | 0.750 | 0.217 | 0.174 | 0.326 | 1.174 |

(\*) S: Significant at $P < 0.05$ ; NS: Non Significant at $P > 0.05$ , informative reportable p value $> 0.05$ if close to ( but does not reach) 0.05 and confirmed by minimum confidence interval (Robert J. Nordness, "Epidemiology
and Biostatistics Secrets", MOSBY ELSEVIER, 1600 John F. Kennedy Boulevard, Suite 1800 Philadelphia, PA 19103-2899, P156, 2006),I: No vaccinated at all; II: No currently vaccinated; III: Currently giving vaccination.

**Figure (1).**
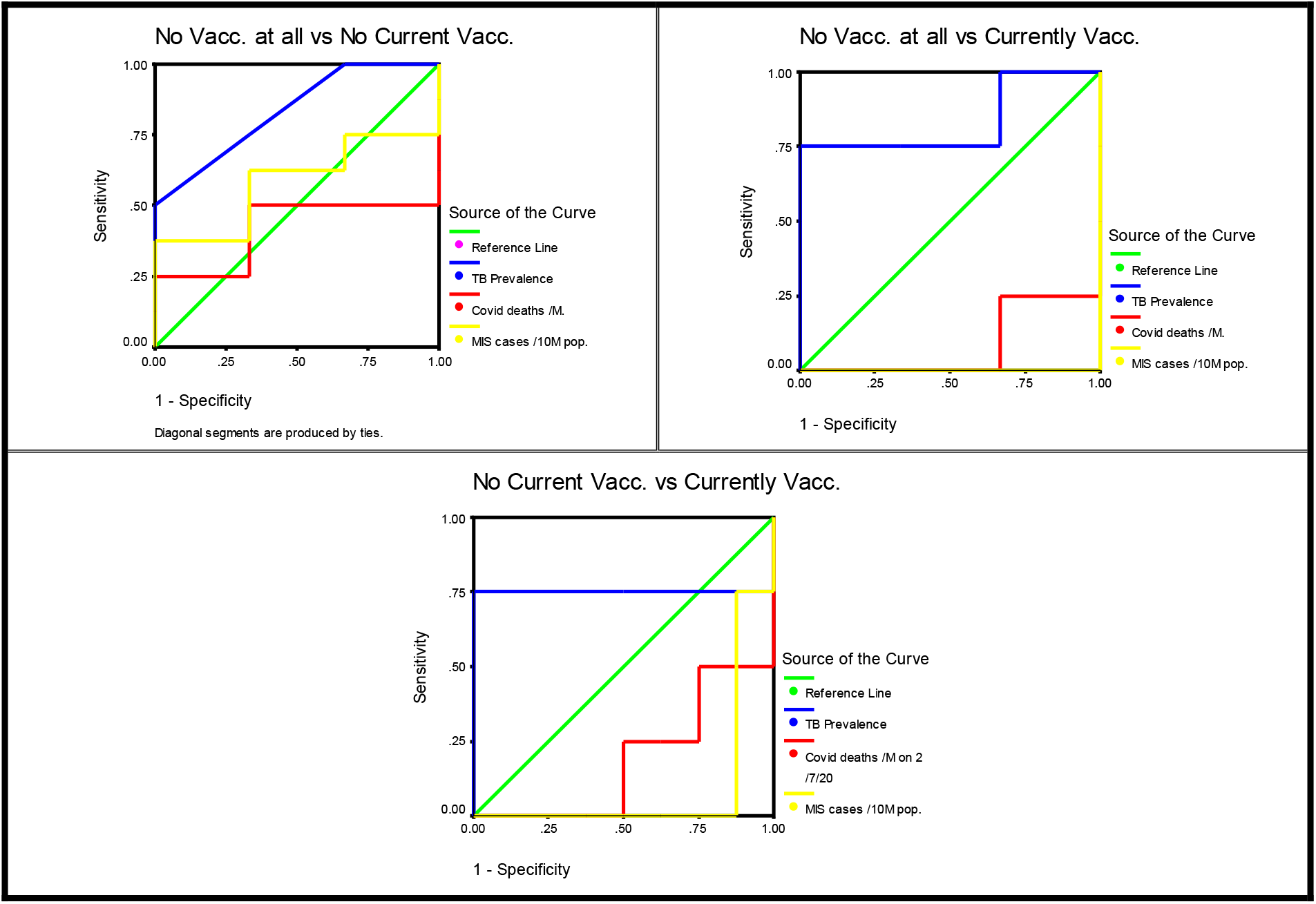
Graphical representation of ROC Curve plots for studied markers in relation with 3 Pair of countries with different BCG status.

Findings are as shown in table 2 and fig. 1: MIS-C no./10 M inhabitants in countries never gave BCG vaccination vs countries currently giving vaccine (pair 2) shows area under ROC-curve equal to 0.000 with a symbiotic significant of 0.000 and (95% CI interval of 0.000-0.000) also MIS-C No/10 M inhabitants in countries not currently give BCG vaccination(ceased mass vaccination programs) vs countries currently give mass vaccination (pair 3) shows area under ROC-curve equal to 0.094 with a symbiotic significant of 0.027and (95% CI interval of 0.000 -0.280).

Important not significant finding is that MIS-C no./10 M inhabitants in countries never gave BCG vaccination vs countries not currently giving vaccine (1^st^ pair) shows area under ROC-curve equal to 0.583 with a symbiotic significant of 0.683 and (95% CI interval of 0.074-0.759). MIS-C no. is a good discriminator in a significant association when countries not currently giving vaccine are in the 3^d^ pair of countries (i.e. with countries currently given BCG vaccine) and not in 1^st^ pair (i.e. with countries never gave national BCG vaccination).

COVID-19 deaths / M inhabitants in countries never gave BCG vaccination vs countries currently giving vaccine (pair 2) show area under ROC-curve equal to 0.083 with a symbiotic informative and reportable value of 0.077 and (95% CI interval of 0.000-0.309 also COVID-19 deaths / M inhabitants in countries not currently giving BCG vaccination vs countries currently giving vaccine(3d pair) show area under ROC-curve equal to 0.188 with a symbiotic informative reportable value of 0.089 and (95% CI interval of 0.000-0.452)

Important finding is the non-significant association of COVID-19 deaths no. /M inhabitants in countries never gave BCG vaccination vs countries not currently give vaccine (1^st^ pair) which shows area under ROC-curve equal to 0.417 with a symbiotic significant of 0.683 and (95% CI interval of 0.078 - 0.755). COVID-19 deaths are a good discriminator in a significant association when countries not currently give vaccine are in third pair of countries (i.e. with countries currently given vaccine) and not in 1^st^ pair (i.e. with countries never gave BCG vaccine in their national programs.

Regarding TB prevalence marker or discriminator the areas under curve were informative and reportable and too generating with the leftover markers in all pairs and were 0.833 with 59% CI of 0.572-0.755 in pair one and 0.833 with 95% CI interval of 0.5-0.500 and 0.750 with 95% CI interval of 0.326-1.174 in pairs 2 and 3 respectively. These signifying inverse relations with COVID-19 mortality and MIS-C no. curves. The informative reportable asymptotic values were 0.102,0.157 and 0.174 for 3 pairs respectively.

In figure (1) TB prevalence marker or discriminator looks to be too generating with the leftover markers in all 3 pairs.

## Discussion

According to ECDC there is lack of understanding with regard to the SARS-CoV-2-induced humoral and cellular immune responses; furthermore, ECDC considered children with this MIS-C for recruitment in research and surveillance studies in addition to descriptive observational studies to further understand the basic clinical and epidemiological parameters associated with this emerging condition.

We suggest that TB and /or BCG might have a role in trained immunity modulation as back ground hypothesis for this study.

Limitations of this study includes but not limited to the following: overlap case definition with Kawasaki disease^25^, retrograde classification of cases, underreporting, under registration, limited no. of countries reported the disease up to 13/6/2020 and limited number of reported cases. Furthermore, this study is done at early time of emergence of this new disease with existence of continuous reporting new cases from different countries in the view of increased awareness of the disease ^39,40,41,42,43, 44^. Also, there was no agreed international nor EU case definition for MIS-C neither specific diagnostic code during study period making the impact of the new syndrome more difficult to be fully assessed.

The statistical negative association between MIS-C and BCG is highly significant between countries which give BCG vaccination versus those not given BCG vaccine. This is statistical association and not necessary means causation although direct beneficial effect to BCG vaccination is suggested. Another mode of protection which might explain this relation in countries already ceased BCG vaccinations which also shows significant negative association is through herd immunity. On the other hand, those with no BCG vaccination are the worst among 3 groups. This negative association also found between BCG practicing and COVID-19 deaths. Similar to BCG, TB prevalence(which reflects latent TB status) shows less significant but informative and reportable association and too generating with the leftover markers. Our findings consolidate previous literatures which suggest influence of BCG and TB prevalence status in lowering COVID-19 mortality31,32,33,34, 35,45. KD emergent cases were back to 1961 when first case identified. In 1967, 50 patients reported by Dr. Tomisaku Kawasaki^46^. Incidence of KD (per 100,000 less than 5-year-olds)^47^ is 308 in Japan in 2014^48^,194.7-217.2 in South Korea in 2014^49,50^, 71.9.03 -116.6 in China in 2010-2014^51^. Recently, greater numbers of countries report incomplete KD cases^52^.

During 1918 pandemic there were no reports for KD or KD like disease condition. Since that time literature research on the impact of Spanish influenza does indicate that the Chinese people survived much better than people in the USA and Europe. There is no evidence of KD like condition in China apart from viral erythema misdiagnosed as scarlet fever reported in some patients in China^53^.

In Europe and USA it seems that the epidemic was started earlier than 1918^54,55, 56, 57, 58,59, 60^. In USA there was widespread of measles epidemic by early October 1917 and continuing into 1918 according to medical reports ^61^. At same time there were increases in scarlet fever and in measles case-fatalities. Measles produced the highest infection rates in 97 years of continuous military surveillance and extreme case-fatality from aggressive bronchopneumonias and other complications^62^. Furthermore, some of the reported deaths were caused by primary pneumonia or empyema without measles^61^. It seems that skin rashes were not reported as a sign of spanish flu in affected persons in USA unlike in China. When KD becomes well known disease, differentiation between KD and measles reported be difficult sometimes ^63^. Furthermore, differentiation between KD or KD like ilness and measles was impossible in 1917-1918 since KD is not yet known, added to that there were no lab facilities to differentiate between 2 diseases. Interstitial bronchopneumonia was used to describe both of 1917 measles and 1918 influenza autopsy findings^61, 64,65^.

Tuberculosis mortality was in decline in the USA since at least the mid-nineteenth century and continued to decline through 1918 during pandemic and later on^66^.

In Japan large-scale influenza epidemics have occurred in the past, including the Spanish flu during 1918–1920, Asian flu during 1957–1958, Hong Kong flu during 1968–1969, and 2009 H1N1 pandemic during 2009–2010. Influenza outbreak in 1957 is caused by (H2N2) and the quite extensive outbreaks in 1962 and 1965 were due to drift variants^67^. Furthermore 13 outbreaks of swine influenza were recognized in Japan from 1978 – 1979^68^. Influenza type B viruses were also associated with increased morbidity in 1982^69^. The 1986-87 influenza epidemic was caused by influenza A(H1N1) variant first isolated in China, Malaysia, Japan, and Singapore^70^. It seems that temporally associated with 4 previous out brakes, three large KD epidemics were recorded in Japan—in 1979, 1982, and in 1986 in addition to the first reported cases by Tomisaku Kawasaki in 1967^46^.

In 1953, Japan was highly endemic with TB. By developments in TB treatments,^71,72^, and giving mass BCG vaccinations since 1948 till now, (boosters were given also before 2003), the mortality rate from TB dropped sharply^72^.

The reported annual incidence rates of KD per 100,000 children aged 0 to 4 years in Japan was 206.2 in 2009 and 239.6 in 2010, the 2010 rate was the highest ever reported before. The influenza epidemic in 2009, (H1N1) was suggested for this increase in incidence^73^.

TB prevalence in Japan was 17 in 2011^74^decreased to 14 in 2018^75^, while KD incidence increased to 308 in 2014. It is possible that decreasing in TB prevalence might be a factor in emerging of KD (in the presence of BCG national vaccination program), while it seems that decreasing TB prevalence might be a factor in KDSS or MIS-C in countries with no current mass BCG vaccinations. This should be confirmed by further studies.

In USA a wide outbreak of KD, apparently the 1^st^ in the USA, occurred in Hawaii in the first half of 1978. Isolates of H1N1 in 50 states in 1977 were reported with new variant of H1N1 reported in few states including Hawaii^76^. During 1984 - 1985, 10 outbreaks of (KD) outbreaks occurred in 10 states and the district of Columbia ^77 .^ According to CDC, during the 1983-1984 influenza season, school and college outbreaks of type A(H1N1) strains began and increased sharply after January 1 and peaked in February^78^, just few months before KD outbreaks.

In South Korea the incidence rates of KD after 2009 influenza pandemic increased from 115.4 in 2009 to 134.4 per 100,000 children in 2011^49,50^ This furtherly increased to 170.9, 194.9 and 194.7 in 2012, 2013 and 2014, respectively^50^.TB prevalence in south Korea was 80 in 2011 decreased to 66 in 2018^79^. Compared to Japan, South Korea have lesser incidence of KD and higher TB prevalence since South Korea start BCG at late times in 70s and stopping boosters in 2007^80^.

In China the start and peak of KD epidemics were in 1979, 1982, and 1986, these out brakes were coincidental to Japan’s for mentioned peaks of KD, furthermore there were seasonal outbreaks between 1987-2010 ^81^. In general, KD incidence is low in China with trends to increase in recent years. In Sichuan Province, the mean incidence (2003-2007) was 7.1^82^and in Hong Kong were 26, 39 and 74 showing increasing manner in 1994, 2000 and 2011 respectively^83^. The incidence is as high as 116.6 in 2014 in Beijing^51^.

The “flu” of 1979(H2 N2) in USA originated in fact in mainland China in early 1977 which was caused by another “new” virus that is not new at all but which circulated widely in the early 1950’s, then vanished with the advent of “Asian” (H2N2) influenza in 1957 ^84^. A new influenza A(H1N1) virus was isolated in China in 1982^85^and another new variant of H1N1 was isolated for first time in China as mentioned before in 1986. TB prevalence in China was 134.27 in 1990 and 137.93 in 2015 per 100,000 persons^86^ and decreased to 61 in 2018^75^.

In Europe, the epidemiology of KD ranges from 3.6 to 15.2/100,000^87^ but (KDSS) has a higher incidence in these countries^13^, compared with Asian countries, the real cause for high incidence of KDSS in western countries is unknown yet.

BCG unlike natural infection gives protection to 50%-60 %^88^ of target population against TB and has immunological heterogeneous effect against some other pathogens^30^. Clinical trials are in progress to establish relations to prevention of SARS-CoV-2 infection.

The temporal associations of MIS-C emergence in different countries with concurrent virus outbrakes and relations of KD or MIS-C emergence to low TB prevalence are of possible concern ad this points to probable but yet not definite causation relation. Furthermore, causation might be back to 1917-1918 pandemic or even before. Furtherly, possible relation of lack of BCG with KDSS or to MIS-C is of further concern too.

### Conclusions

This study shows a significant relation between BCG and both of incidence of MIS-C and COVID – 19 deaths, also between TB prevalence and both of incidence of MIS-C and COVID-19 deaths.

### Recommendations

Controlled clinical trials are required to confirm causation of TB prevalence and BCG status to MIS-C or to Kawasaki like spectrum of diseases in general.

### Ethics and dissemination

Ethical permission is not necessary as this study analyzed publically published data and patients were not involved.

There is no conflict of interest.

There is no funding received.

## Data Availability

All relevant data are available on requist

## Acknowledgment

I am deeply grateful to Emeritus Professor Abdulkhaleq Abduljabbar Ali Ghalib Al-Naqeeb, Ph.D. in the Philosophy of Statistical Sciences at the Medical & Health Technology college, Baghdad-Iraq, for his assistance and support in data analysis, interpret

**Abbreviations**
CDC: Centre for Disease Control
COVID-19: coronavirus disease of 2019
ECDC: European Centre for Disease Prevention and Control
KD: Kawasaki disease
MIS-C: Multisystem inflammatory syndrome in children
PIMS-TS: Pediatric inflammatory multisystem syndrome temporally associated with SARS-CoV-2 infection in children
SARS-CoV-2: Severe acute respiratory syndrome coronavirus 2

